# Determining the importance of risk factors for the occurrence of temporomandibular disorders in the population and among exposed individuals

**DOI:** 10.1101/2024.01.31.24302055

**Authors:** James R. Miller, Zachary J. Hass

## Abstract

There are many case-control (Ca-Co) studies in the literature on the importance of risk factors for the occurrence of temporomandibular disorders (TMD). These studies typically report the adjusted odds ratio (OR) for each risk factor being studied. This paper presents other epidemiological measurements for evaluating the importance of risk factors for the occurrence of TMD. These measurements include the population attributable risk percent (PAR%) and the attributable risk percent (AR%). The AR% for parafunctional habits, facial trauma, and orthopedic instability were estimated to be 86%, 80%, and 60%, respectively, while the corresponding PAR% were estimated to be 60%, 38%, and 7%. PAR% underestimates the importance of a risk factor for the occurrence of TMD among individuals exposed to the risk factor. The attributable risk percent (AR%) is an appropriate epidemiological measurement to evaluate the importance of a risk factor for TMD among individuals exposed to the risk factor.

## Introduction

The cumulative incidence of painful TMD in the population has been reported to be about 4%. [1] This, in combination with the fact that most risk factors for TMD cannot be randomly assigned to participants in studies, helps explain why much of the research on risk factors for the occurrence of TMD are Ca-Co studies. Ca-Co studies often report adjusted odds ratios (OR) for each risk factor being studied to indicate the strength of association of a risk factor with TMD. However, other epidemiological measurements that are useful for determining the importance of risk factors for the occurrence of TMD, both in the population and among exposed individuals, are not routinely reported. These include the population attributable risk percent (PAR%) and attributable risk percent (AR%).

Research on risk factors for the occurrence of TMD may be conducted on samples from the population and the results from such research are frequently reported from the perspective of the importance of the risk factors for the occurrence of TMD in the population. However, clinicians are generally more interested in the importance of risk factors for the development of TMD among individuals exposed to one or more risk factors. The importance of risk factors for the occurrence of TMD needs to be considered from these two perspectives, the occurrence in the population and the occurrence among individuals exposed to one or more risk factors.

The importance of a risk factor for disease in the population is not necessarily the same as the importance of the risk factor for disease among individuals exposed to that risk factor. Fortunately, population-based studies on risk factors for TMD often provide the data needed to calculate epidemiological measurements that are useful for evaluating both the importance of a risk factor for disease in the population and among exposed individuals.

The objectives of this study are to present epidemiological measurements for evaluating the importance of risk factors for the occurrence of TMD and illustrate their calculation and interpretation through examples that evaluate the importance of (1) one risk factor for painful TMD, (2) three risk factors for TMD, and (3) eight combinations of these three risk factors for TMD. The definitions of the epidemiological measurements presented in this article and the formulas for calculating these measurements are provided in the Appendix or in the referenced textbooks. [2] [3]

## Materials and Methods

Epidemiological measurements, as they pertain to evaluating the importance of risk factors for the occurrence of TMD, are calculated using standard formulas. The calculated measurements include the attributable risk percent (AR%) and the population attributable risk percent (PAR%), and where possible, the attributable risk (AR) and the population attributable risk (PAR). These calculations require the input of other epidemiological measurements. These input measurements include the cumulative incidence among the exposed (Ie), the cumulative incidence among the non-exposed (Io), the relative risk (RR) as approximated by the odds ratio (OR), and the prevalence of risk factors in the population (pe). In this study, values for these input measurements are based on values found in the literature.

The first example examines the importance of one dichotomous risk factor for the occurrence of painful TMD (Table 1). The risk factor is “any injury”, with individuals being categorized as either exposed or non-exposed. The values for the input variables, the odds ratio (OR) for the association between this risk factor and painful TMD and the prevalence of this risk factor in the population (pe), were obtained from a large community-based Ca-Co study. [4] The cumulative incidence of painful TMD in the population being studied (It) was also available [1], which allowed the calculation of the values for the input variables cumulative incidence among the exposed (Ie) and cumulative incidence among the non-exposed (Io). The attributable risk percent (AR%), population attributable risk percent (PAR%), attributable risk (AR), and the population attributable risk (PAR) were calculated using the values for the input variables Ie, Io, RR, and pe (Table 1).

**Table 1.**
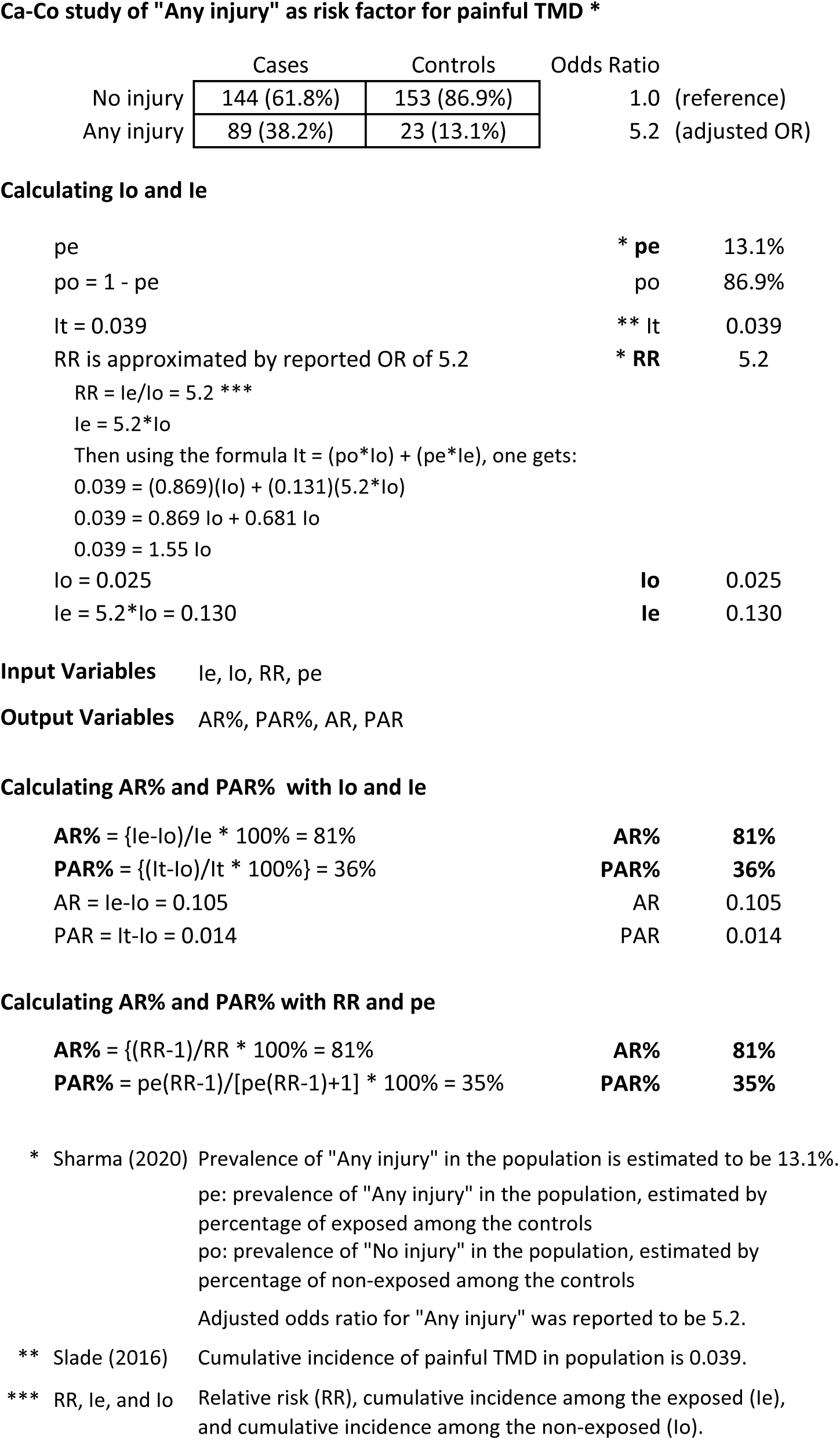
AR% and PAR% for a single risk factor for painful TMD.

**Ca-Co study of "Any injury" as risk factor for painful TMD \***
|  | Cases | Controls | Odds Ratio |
| --- | --- | --- | --- |
| No injury | 144 (61.8%) | 153 (86.9%) | 1.0 (reference) |
| Any injury | 89 (38.2%) | 23 (13.1%) | 5.2 (adjusted OR) |

**Calculating lo and le**
|  |  |  |
| --- | --- | --- |
| pe | * pe | 13.1% |
| po = 1 - pe | po | 86.9% |
| It = 0.039 | ** It | 0.039 |
| RR is approximated by reported OR of 5.2 | * RR | 5.2 |
| RR = le/lo = 5.2 *** |  |  |
| le = 5.2 * lo |  |  |
| Then using the formula It = (po * lo) + (pe * le), one gets: |  |  |
| 0.039 = (0.869)(lo) + (0.131)(5.2 * lo) |  |  |
| 0.039 = 0.869 lo + 0.681 lo |  |  |
| 0.039 = 1.55 lo |  |  |
| lo = 0.025 | lo | 0.025 |
| le = 5.2 * lo = 0.130 | le | 0.130 |

**Calculating AR% and PAR% with lo and le**
|  |  |  |
| --- | --- | --- |
| AR% = (le-lo)/le * 100% = 81% | AR% | 81% |
| PAR% = ((It-lo)/It * 100%) = 36% | PAR% | 36% |
| AR = le-lo = 0.105 | AR | 0.105 |
| PAR = It-lo = 0.014 | PAR | 0.014 |

**Calculating AR% and PAR% with RR and pe**
|  |  |  |
| --- | --- | --- |
| AR% = {(RR-1)/RR * 100% = 81% | AR% | 81% |
| PAR% = pe(RR-1)/[pe(RR-1)+1] * 100% = 35% | PAR% | 35% |

\* Sharma (2020) Prevalence of "Any injury" in the population is estimated to be 13.1%.
pe: prevalence of "Any injury" in the population, estimated by percentage of exposed among the controls
po: prevalence of "No injury" in the population, estimated by percentage of non-exposed among the controls
Adjusted odds ratio for "Any injury" was reported to be 5.2.
\*\* Slade (2016) Cumulative incidence of painful TMD in population is 0.039.
\*\*\* RR, le, and lo Relative risk (RR), cumulative incidence among the exposed (le), and cumulative incidence among the non-exposed (lo).

The next example examines the importance of multiple risk factors [5] [6] [7] for the occurrence of TMD, either muscle disorders, temporomandibular joint (TMJ) disorders, or a combination of the two (Table 2). The risk factors are parafunctional habits, facial trauma, and orthopedic instability. The values for the input variables, the odds ratio (OR) and the prevalence in the population (pe) for each of these risk factors, were based on a range of values from studies reported in the literature. [4] [8] [9] [10] [11] [12] [13] [14] [15] The attributable risk percent (AR%) and the population attributable risk percent (PAR%) for each of the three risk factors in this example were calculated using these input values. These calculations were made using the various assumptions listed in Table 2. This is an example of a type of Ca-Co study frequently found in the literature, which typically reports the adjusted odds ratio for each risk factor being investigated.

**Table 2.** Multiple risk factors for TMD.

**Adjusted OR, AR% and PAR% for three risk factors for TMD #**
|  |  | OR * | AR% ** | pe * | PAR% *** |
| --- | --- | --- | --- | --- | --- |
| **PAF** | non-exp | 1.0 |  |  |  |
|  | exposed | **7.0** | 86% | 25% | 60% |
| **TRA** | non-exp | 1.0 |  |  |  |
|  | exposed | **5.0** | 80% | 15% | 38% |
| **OIS** | non-exp | 1.0 |  |  |  |
|  | exposed | **2.5** | 60% | 5% | 7% |

The third example examines the importance of eight combinations of risk factors for the occurrence of TMD (Tables 3 and 4). The three risk factors and the values for the input variables used in the previous example were used in this example. The prevalence for these three risk factors were used to estimate the prevalence in the population for each of the eight combinations in this example (Table 3). The odds ratios for each risk factor in a combination were used to calculate the OR for that combination (Table 4). The combination prevalence and the combination OR for each combination were used to calculate the combination AR% and combination PAR% for each combination (Table 4).

**Table 3.** Prevalence of combinations of risk factors in the population.

**Prevalence of each of the three risk factors in the population** Taken from Table 2.
|  | **PAF** | **TRA** | **OIS** |
| --- | --- | --- | --- |
| prevalence of non-exposure (po) | 75% | 85% | 95% |
| prevalence of exposure (pe) | 25% | 15% | 5% |

**Prevalence of combinations of these three risk factors in the population** All possible combinations of the above three risk factors are listed.
|  | Prevalence of non-exposure and<br>exposure for each risk factor |  |  | **Combination**<br>**prevalence <sup>^</sup>** |
| --- | --- | --- | --- | --- |
|  | **PAF** | **TRA** | **OIS** |  |
| non-exposed to all | 75% | 85% | 95% | 60.6% |
| exposed PAF | 25% | 85% | 95% | 20.2% |
| exposed TRA | 75% | 15% | 95% | 10.7% |
| exposed OIS | 75% | 85% | 5% | 3.2% |
| exposed PAF and TRA | 25% | 15% | 95% | 3.6% |
| exposed PAF and OIS | 25% | 85% | 5% | 1.1% |
| exposed TRA and OIS | 75% | 15% | 5% | 0.6% |
| exposed PAF, TRA, and OIS | 25% | 15% | 5% | 0.2% |

<sup>^</sup> The **combination prevalence** is calculated by multiplying the prevalences for all risk factors in the combination.
Assumptions:
- (1) No interactions between risk factors, and - (2) Risk factors are statistically independent

**Table 4.** Combinations of risk factors for TMD.

|  | PAF | TRA | OIS | Combination | Combination |  | Combination | Combination |
| --- | --- | --- | --- | --- | --- | --- | --- | --- |
|  | OR | OR | OR | OR * | AR% ** |  | prevalence ^ | PAR% *** |
| non-exposed to all | 1.0 | 1.0 | 1.0 | 1.0 | 0% | po | 60.6% | 0% |
| exposed PAF | 7.0 | 1.0 | 1.0 | 7.0 | 86% | pe | 20.2% | 28% |
| exposed TRA | 1.0 | 5.0 | 1.0 | 5.0 | 80% | pe | 10.7% | 10% |
| exposed OIS | 1.0 | 1.0 | 2.5 | 2.5 | 60% | pe | 3.2% | 1% |
| exposed PAF and TRA | 7.0 | 5.0 | 1.0 | 35.0 | 97% | pe | 3.6% | 28% |
| exposed PAF and OIS | 7.0 | 1.0 | 2.5 | 17.5 | 94% | pe | 1.1% | 4% |
| exposed TRA and OIS | 1.0 | 5.0 | 2.5 | 12.5 | 92% | pe | 0.6% | 2% |
| exposed PAF, TRA, and OIS | 7.0 | 5.0 | 2.5 | 87.5 | 99% | pe | 0.2% | 4% |

\* The **combination OR** for each combination of risk factors is calculated by multiplying the ORs for all three risk factors in the combination. The combination OR approximates the **combination RR**. The combination RR is an input variable for the combination AR% and combination PAR%.
^ The **combination prevalence** are taken from Table 3. The combination prevalence is an input variable for the combination PAR%.
\*\* The **combination AR%** for each combination of risk factors is calculated by using the combination OR as an approximation for the combination RR and then by using the formula:
\*\*\* The **combination PAR%** for each combination of risk factors is calculated by using the combination OR as an approximation for the combination RR and the combination prevalence and then by using the formula:
$$\text{pe}(\text{combination RR}-1)/[\text{sum}\{\text{pe}(\text{combination RR}-1)\}+1] * 100\%.$$ Note: This formula is used to calculate the PAR% for each mutually exclusive combination. The aggregate PAR% is the sum of these.

## Results

The results in these examples all pertain to the importance of risk factors for the occurrence of TMD. The epidemiological measurements AR%, and PAR%, and where possible AR and PAR, were calculated for these examples. The input variables for calculating these measurements included the Ie, Io, RR, and pe.

In the first example, AR%, PAR%, AR, and PAR were calculated for the risk factor “any injury” to evaluate the importance of this risk for the occurrence of painful TMD (Table 1). The cumulative incidence of painful TMD in this population (It) was reported by Slade to be 0.039, or approximately 4%. [1] In 2020, Sharma using data from the same study as Slade investigated “any injury” as a risk factor for painful TMD and reported the adjusted odds ratio (OR) for “any injury” to be 5.2. [4] This OR was used to approximate the relative risk (RR). Sharma also reported that 13.1% of the controls were exposed to “any injury”, which provided an estimate of the prevalence of “any injury” in the population (pe). [4] It, RR, and pe allowed the calculation of the cumulative incidence of TMD among the exposed (Ie) and among the non-exposed (Io). Then, Ie, Io, RR, and pe were used to calculate the AR%, PAR%, AR, and PAR for “any injury”, which produced the following values, 81%, 36%, 0.105, and 0.014.

In the second example, AR% and PAR% were calculated for multiple risk factors to evaluate the importance of these risk factors for the occurrence of TMD (Table 2). The odds ratios selected for the three risk factors, parafunctional habits, facial trauma, and orthopedic instability were 7.0, 5.0, and 2.5. These OR were used to approximate the RR for each risk factor. The prevalence in the population selected for these three risk factors were 25%, 15%, and 5%. Using these RR and pe, the AR% for the three risk factors were calculated to be 86%, 80%, and 60%, and the PAR% were calculated to be 60%, 38%, and 7%.

In the third example, AR% and PAR% were calculated for combinations of risk factors to evaluate the importance of these combinations for the occurrence of TMD (Table 3 and 4). The three risk factors from the previous example were used in this example to produce eight mutually exclusive combinations of risk factors. The OR for each combination was calculated using the OR for each risk factor in the combination (Table 4). The odds ratios for the combinations of risk factors, with exposure to at least one risk factor, ranged from 2.5 to 87.5. The prevalence of each combination in the population was calculated using the population prevalence of each risk factor in the combination (Table 3). The combination RR, as approximated by the combination OR, and the combination pe were the input values used to calculate the combination AR% and combination PAR%. The AR% for the combinations of risk factors, with exposure to at least one risk factor, ranged from 60% to 99%. The PAR% for the combinations of risk factors, with exposure to at least one risk factor, ranged from 1% to 28%.

## Discussion

The objectives of this study were to present epidemiological measurements for evaluating the importance of risk factors for the occurrence of TMD. The importance of a risk factor for the occurrence of TMD depends on whether one is primarily interested in the importance of the risk factor for the occurrence of TMD in the population, among individuals exposed to the risk factor, or for an individual exposed to the risk factor. The PAR% is useful for evaluating the portion of TMD cases in the population attributable to a risk factor. The AR% is useful for evaluating the portion of TMD cases in individuals exposed to a risk factor that is attributable to the risk factor. The AR% for a risk factor is also useful for evaluating the likelihood that a TMD in an individual is attributable to the risk factor.

For clinicians who treat individual patients, AR% is a useful epidemiological measurement because it indicates the likelihood that a TMD in an individual is attributable to a risk factor. The adjusted OR for risk factors are often reported in studies of risk factors for the occurrence of TMD, but AR% are not routinely reported. However, it is easy to use an adjusted OR to calculate the corresponding AR% by using the OR as an approximation of the RR and then using the formula AR% = (RR-1)/RR *100%. [2] This formula was used to calculate the AR% for the three risk factors in the second example (Table 2). This approach could also be used to calculate AR% for other purported risk factors for TMD, such as systemic diseases and psychological disorders.

The calculation and interpretation of PAR% for multiple risk factors is more complex. [16, 17] PAR% depends on both the prevalence of a risk factor in the population and the RR associated with that risk factor. Further, because there are many risk factors for TMD and an individual can have more than one risk factor at a time, the PAR% for each risk factor included in a study of multiple risk factors do not sum to the PAR% for the risk factors in aggregate. [16] This is illustrated with the PAR% for the three risk factors investigated in the second example (Table 2), which if summed would produce an inaccurate value for the PAR% for the three risk factors in aggregate. Using an appropriate formula produces the correct PAR% for the three risk factors in aggregate of 77%. [16] Since an individual can have only one combination of risk factors at a time, the PAR% for the eight combinations of risk factors in the last example (Table 4) do sum to the correct PAR% of 77% for the three risk factors in aggregate. [17] Understanding PAR% for multiple risk factors can be complex and is introduced here mostly for the sake of completeness and for those who might be interested.

A concept that is particularly important is that a risk factor can account for a large portion of cases among those exposed to the risk factor, while at the same time accounting for only a small portion of cases in the population. This is illustrated by the AR% and PAR% for the risk factor orthopedic instability (OIS) in the second example (Table 2). According to these data, OIS accounts for 60% of TMD among those exposed to the risk factor, while accounting for only 7% of TMD in the population. For an individual with OIS who develops TMD, this means that the likelihood that the TMD in this individual is due to exposure to OIS is 60%, not 7%. This example illustrates that a risk factor can be important for the occurrence of TMD among individuals exposed to a risk factor (AR%), while at the same time the risk factor is relatively unimportant for the occurrence of TMD in the entire population (PAR%). Further, this example illustrates that AR% is an appropriate measurement to evaluate the importance of a risk factor for the occurrence of TMD among individuals exposed to the risk factor.

This concept is related to another important concept presented in some articles using logistic regression to evaluate risk factors for TMD. [10] [14] These articles use statistical models to calculate the adjusted odds ratio for a risk factor and to approximate the portion of variation of TMD in a population that is explained by the risk factor using Pseudo R^2 measures. When a risk factor is evaluated in this manner, a risk factor can have a significant odds ratio, while at the same time explain only a small portion of the variation of TMD in a population. However, a small variation of TMD in a population explained by a risk factor does not necessarily mean a risk factor is clinically unimportant. A risk factor can be clinically important for the occurrence of TMD among exposed individuals, while at the same time explain only a small portion of the variation of TMD in a population, similar to the situation where orthopedic instability (OIS) has a large AR%, while at the same time having a small PAR%.

The eight possible combinations of three risk factors for TMD are presented in the third example, along with the calculations for the OR, AR%, and PAR% for these combinations (Table 4). The OR and AR% for a combination of risk factors can be quite large. A large AR% for a combination would indicate a correspondingly large likelihood that the TMD in an individual exposed to the combination is due to this combination. This type of study is generally not attempted for investigating risk factors for TMD because of the challenge of recruiting enough participants for the rarer combinations.

Prevention of TMD by mitigating the influence of one or more risk factors is not the primary focus of this article. Evaluating whether or not to try to prevent TMD by mitigating the influence of one or more risk factors is a complex topic. [18] A complete evaluation of whether to institute a preventive intervention in an exposed individual is best made knowing both the AR% and AR. Unfortunately, AR is often not known because the cumulative incidence of disease among the exposed (Ie) and non-exposed (Io) cannot be determined with data from the typical Ca-Co study that is not population-based. In general, before instituting a preventive intervention, one needs to consider the necessity and practicality of screening a population to identify individuals exposed to a risk factor and the availability, effectiveness, safety, cost, and side effects associated with a particular preventive intervention that might be used to mitigate the influence of the risk factor.

For example, for “any injury”, where the cumulative incidence of painful TMD in the population (It) is known, the Ie and Io could be calculated and were 0.130 and 0.025. These values indicate the risk of developing a painful TMD among the exposed and non-exposed (Table 1). Using these incidence data, AR% was calculated to be 81% and AR was calculated to be 0.105. The AR% indicates that 81% of painful TMD cases among exposed individuals could be prevented per year. However, the AR indicates the actual number of cases that could be prevented would only be about 11 cases per 100 exposed individuals per year once all 100 exposed individuals had been provided with the preventive intervention. For an exposed individual, the AR% means a preventive intervention could reduce the risk of an exposed individual developing a painful TMD by 81%, but the AR means the absolute reduction in risk would only be 0.105, a reduction from 0.130 to 0.025. If one only knows the AR%, one might attempt prevention in an individual exposed to “any injury”. However, if one also knows the AR, one might want to reconsider, especially if the proposed preventive intervention is costly or risky.

The second and third examples (Tables 2-4) looked at three risk factors, including parafunctional habits, facial trauma, and orthopedic instability. The values for the input variables, the odds ratio (OR) and prevalence in the population (pe) for these three variables, were based on a range of values in the literature. It would have been best to have consensus point estimates from meta-analyses for the values for the input variables used in the last two examples (Tables 2-4). However, given the variation of definitions for risk factors and TMD in the available research, meta-analyses are currently of limited use for identifying consensus point estimates for the odds ratios for the association between risk factors and TMD. [15]

There were certain considerations when selecting the values for the OR and pe for these risk factors. Some of the odds ratios for the association between parafunctional habits and TMD that have been reported in the literature are large and are likely due, in part, to recall bias. [1] [11] Thus, a conservative value for the OR was used for parafunctional habits. With respect to orthopedic instability (OIS), there is surprisingly little research on OIS as a risk factor for TMD, per se, even though it is considered an important risk factor for TMD by Okeson. [5] [6]

It would be best if epidemiological studies on risk factors for TMD were able to use a validated “instrument” to measure orthopedic instability (OIS). Direct measurements of OIS in epidemiological studies on risk factors for TMD are currently not readily available. However, what are available are epidemiological studies that measure static and dynamic malocclusions as possible risk factors for the occurrence of TMD. How good malocclusions are as proxy measurements for OIS is unknown. Regardless, in this paper, the odds ratios and prevalence of malocclusions in the population from several studies on malocclusions and TMD were used as the basis for selecting the values for the odds ratio (OR) and prevalence in the population (pe) for the risk factor orthopedic instability (OIS). [10] [11] [12] [14] [15]

Since TMD are relatively rare [1], the use of OR to approximate RR is reasonable, although a limitation. [19] However, there were some concerns that the values chosen for the input variables OR and pe for the three risk factors in the second and third examples might not have been the most appropriate. To allay these concerns, calculations were repeated using a reasonable range of values for the OR and pe to calculate AR% and PAR%. The results using these range of values were consistent with the results reported in this paper. Thus, the potential limitation of selecting values from a range of values in the literature, for the input variables OR and pe for the second and third examples (Tables 2-4), would seem to be minimal.

In the first example that evaluated the relationship of the risk factor “any injury” to painful TMD, these TMD included myalgia and/or arthralgia. In the second and third examples, the TMD in studies in the literature from which input values were selected included muscle disorders and/or joint disorders. Since many individuals have combinations of muscle and joint disorders [11] [12] [13] [15], evaluating how risk factors influence the occurrence of specific muscle or joint disorders is difficult. Even studies that strive to look at these disorders separately end up with many participants that have a combination of muscle and joint disorders [11] [12] [15]. Looking at combinations of TMD in this study is a limitation, but a limitation that is likely present in many, if not most, research on risk factors for the occurrence of TMD.

Ultimately, the above-mentioned considerations and limitations do not change the main points of this article, that the importance of a risk factor for TMD in the population is not the same as its importance for TMD among individuals exposed to the risk factor, that the importance of a risk factor for TMD among individuals exposed to the risk factor is greater than its importance for TMD in the population, and that the attributable risk percent (AR%) is an appropriate epidemiological measurement to evaluate the importance of a risk factor for TMD among individuals exposed to the risk factor.

## Conclusions

The epidemiological measurements presented in this article should be of particular interest to clinicians who previously may not have appreciated how these measurements are applicable to individual patients in clinical practice. This article illustrates that judging the importance of risk factors for the occurrence of TMD is complicated but can be facilitated by using appropriate epidemiological measurements.

The main points of this article are:

1. The importance of a risk factor for TMD in the population is not the same as its importance for TMD among individuals exposed to the risk factor,
2. The importance of a risk factor for TMD among individuals exposed to the risk factor is greater than its importance for TMD in the population, and
3. The attributable risk percent (AR%) is an appropriate epidemiological measurement to evaluate the importance of a risk factor for TMD among individuals exposed to the risk factor.

## Data Availability

All data used in the submitted manuscript to calculate the various epidemiological measurements are available within the manuscript or in the cited references.

---

**Appendix.** Epidemiological measurements of excess risk of disease for a dichotomous risk factor *.

|  | Population | Exposed Individuals | Exposed Individual |
| --- | --- | --- | --- |
| **Relative Risk**<br>$I_e / I_o$ | | The ratio of the amount of disease among the exposed to the amount of disease among the non-exposed. ** | |
| **Attributable Risk %**<br>$\{ (I_e - I_o) / I_e \} * 100\%$ | | Percent of disease in individuals exposed to a risk factor that is attributable to that exposure. | Likelihood that a disease in a person exposed to a risk factor is attributable to that exposure. |
| **Pop. Attributable Risk %**<br>$\{ (I_t - I_o) / I_t \} * 100\%$ | Percent of disease in the population that is attributable to exposure to a risk factor. | | |
| **Attributable Risk**<br>$I_e - I_o$ | | Amount of disease in individuals exposed to a risk factor that is attributable to that exposure. | |
| **Pop. Attributable Risk**<br>$I_t - I_o$ | Amount of disease in the population that is attributable to exposure to a risk factor. | | |

\* Adapted from:
(1) T.D. Koepsell and N.S. Weiss, "Measures of Excess Risk", in *Epidemiology Methods*, New York, NY, USA: Oxford University Press, 2003, pp. 196-214.
(2) C. H. Hennekens and J. E. Buring, "Measures of Disease Frequency and Association," in *Epidemiology in Medicine*, Boston, Little, Brown and Company, 1987, pp. 54-98.
\*\* The term "amount of disease" is referring to the cumulative incidence of disease.

## Bibliography

[1] G. D. Slade, R. Ohrbach and, et al., "Painful temporomandibular disorders: decade of discovery from OPPERA studies," Journal of Dental Research, vol. 95, no. 10, pp. 1084-1092, 2016.

[2] T. D. Koepsell and N. S. Weiss, "Measures of Excess Risk," in Epidemiologic Methods: Studying the Occurrence of Illness, New York, Oxford University Press, 2003, pp. 196-214.

[3] C. H. Hennekens and J. E. Buring, "Measures of Disease Frequency and Associàon," in Epidemiology in Medicine, Boston, Lible, Brown and Company, 1987, pp. 54–98.

[4] S. Sharma, R. Ohrbach, R. B. Fillingim, J. D. Greenspan and G. Slade, "Pain sensìvity modifies risk of injury-related temporormandibular disorder," Jounal of Dental Research, vol. 99, no. 5, pp. 530–536, 2020.

[5] J. Okeson, F. B. Porto, B. D. Furquim, D. Feu, F. Sato and L. Cardinal, "An interview with Jeffrey Okeson," Dental Press J Orthod, vol. 23, no. 6, pp. 30-39, 2018.

[6] J. P. Okeson, Management of Temporomandibular Disorders and Occlusion, St. Louis: Elsevier, 2020.

[7] D. Manfredini, "Èopathogenesis of disk displacement of the temporomanidbular joint: a review of the mechanisms," Indian Journal of Dental Research, vol. 20, no. 2, pp. 212–221, 2009.

[8] P. Wetselaar and et al., "The prevalence of awake bruxism and sleep bruxisum in the Dutch adolescent populàon," Journal Oral Rehabilita@on, vol. 48, pp. 143–149, 2021.

[9] S. Azami-Aghdash and et al., "Prevalence, èology, and types of dental trauma in children and adolescents: systemàc review and meta-analysis," Med J Islam Repub Iran, vol. 29, p. 234, 2015.

[10] A. G. Pullinger, D. A. Seligman and J. A. Gornbein, "A multiple logistic regression analysis of the risk and relàve odds of temporomandibular disorders as a function of common occlusal features," Journal of Dental Research, vol. 72, no. 6, pp. 968–979, 1993.

[11] J. R. Miller, J. A. Burgess and C. W. Critchlow, "Associàon between mandibular retrognathia and TMJ disorders in adult females," J Public Health Dent, vol. 64, no. 3, pp. 157–163, 2004.

[12] J. R. Miller, L. Mancl and C. Critchlow, "Severe retrognathia as a risk factor for recent onset painful TMJ disorders among adult females," Journal of Orthodon@cs, vol. 32, pp. 249–256, 2005.

[13] R. Ohrbach and, et al., "Clinical findings and pain symptoms as potential risk factos for chronic TMD: descriptive data and empirically identified domains from the OPPERA case-control study," Journal Pain, vol. 12, no. 11 (November), Suppl. 3, pp. T27-T45, 2011.

[14] D. Manfredini, G. Perineh and L. Guarda-Nardini, "Dental malocclusion in not related to temporomandibular joint clicking; a logistic regression analysis in a pàent populàon," Angle Orthodon@st, vol. 84, no. 2, pp. 310–315, 2014.

[15] C. L. Cruz, K. C. Lee, J. H. Park and A. I. Zavras, "Malocclusion characteristics as risk factors for temporomandibular disorders: lessons learned from meta-analysis," Journal of Oral Diseases, vol. Article ID 302646, pp. 1-10, 2015.

[16] P. Bruzzi, S. B. Green, D. P. Byar, L. A. Brinton and C. Schairer, "Estimàng the populàon abributable risk for multiple risk factors using case-control data," American Journal of Epidemiology, vol. 122, no. 5, pp. 904–914, 1985.

[17] M. Chang, R. A. Hahn, S. M. Teutsch and L. C. Hutwagner, "Multiple risk factors and populàon abributable risk for ischemic heart disease mortality in the United States, 1971-1992," Journal of Clinical Epidemiology, vol. 54, pp. 634-644, 2001.

[18] J. Miller and L. Mancl, "Risk factors for the occurrence and prevention of temporomandibular joint and muscle disorders: lessons from 2 recent studies," Am J Orthod Dentofacial Orthop, vol. 134, pp. 537–542, 2008.

[19] H. T. O. Davies, I. K. Crombie and M. Tavakoli, "When can odds ràos mislead?," BMJ, vol. 316, no. 28 March, pp. 989-991, 1998.

